# Galectin antagonist use in mild cases of SARS-CoV-2; pilot feasibility randomised, open label, controlled trial

**DOI:** 10.1101/2020.12.03.20238840

**Authors:** Alben Sigamani, Samarth Shetty, Madhavi, Mathu Ruthra, Sudhishma, Anup Chugani, Hana Chen-Walden, ThomasKutty, David Platt

## Abstract

**Importance:** Novel SARS-CoV-2 virus has infected nearly half a billion people across the world and is highly contagious. There is a need for a novel mechanism to block viral entry and stop its replication.

**Background:** Spike protein N terminal domain (NTD) of the novel SARS-CoV-2 is essential for viral entry and replication in human cell. Thus the S1 NTD of human coronavirus family, which is similar to a galectin binding site - human galactose binding lectins, is a potential novel target for early treatment in COVID-19.

**Objectives:** To study the feasibility of performing a definitive trial of using galectin antagonist – Prolectin-M as treatment for mild, symptomatic, rRT-PCR positive, COVID-19.

**Main outcomes and measures:** Cycle threshold (Ct) value is number of cycles needed to express fluorescence, on real time reverse transcriptase polymerase chain reaction. Ct values expressed for RNA polymerase (Rd/RP) gene +Nucleocapsid gene and the small envelope (E) genes determine infectivity of the individual. A digital droplet PCR based estimation of the Nucleocapid genes (N1+N2) in absolute copies/μL determines active viral replication.

**Design and intervention:** Pilot Feasibility Randomised Controlled Open-Label, parallel arm, study. Oral tablets of Prolectin-M were administered along with the best practice, Standard of Care (SoC) and compared against SoC. Voluntarily, consenting individuals, age >18 years, and able to provide frequent nasopharyngeal and oropharyngeal swabs were randomly allocated by REDCap software.

The intervention, Prolectin-M was administered as a multi dose regime of 4 gram tablets. Each tablet contained 2 grams of (1-6)-Alpha-D-mannopyranosil mixed with 2 grams of dietary fibre. Each participant took a single chewable tablet every hour, to a maximum of 10 hours in a day. Tablets were administered only during the daytime, for total of 5 days.

**Results:** This pilot trial demonstrated the feasibility to recruit and randomize participants. By day 7, following treatment with Prolectin-M, Ct value of Rd/Rp + N gene increased by16.41 points, 95% (CI – 0.3527 to 32.48, p=0.047). Similarly, small envelope (E) gene also increased by 17.75 points (95% CI;-0.1321 to 35.63, p = 0.05). The expression of N1, N2 genes went below detectable thresholds by day 3 (Mann Whitney U = 0.000, p<0.029).

rRT-PCR testing done in the clinic on day 1, 7, and 14 had 3 participants (60%) turn negative by day 7 and all turned negative by day 14 and stayed negative until day 28. In the SoC group 2 participants had zero detectable viral loads at baseline, 2 participants tested negative on day 14, and the last participant tested remained positive on day 28. There were no serious adverse events, and all participants were clinically asymptomatic before day 28 with reactive immunoglobulin G (IgG).

**Trial relevance:** This pilot study proves that it is feasible and safe to perform a trial using a Galectin antagonist in COVID-19. This is a novel mechanism for blocking viral entry and its subsequent replication.

**Trial Registration:** Clinical Trials.gov identifier NCT04512027; CTRI ref. CTRI/2020/09/027833

## Introduction

SARS-CoV-2 has infected over 40 million people worldwide and is responsible for over 1.1 million deaths.^1^ Several treatments have been authorized by regulators around the globe and none of them have been able to lower its infectivity.^2^ SARS-CoV-2, a new member of the coronaviruses known to infect humans,^3^ is a single-stranded RNA-enveloped virus, with a large number of glycosylated S proteins covering its surface.^4^ These proteins mediate viral cell entry following a S protein binding on host cell surface.^5^

During viral infection, the target cells activate the S protein, by cleaving it into S1 and S2 subunits.^6^ The S1 subunit is further divided into a N-terminal domain (NTD) and a C-terminal domain (CTD). The CTD is known to bind the human angiotensin converting enzyme-2 (ACE-2) receptors while NTD seeks gangliosides on cell surface to stabilize cell adhesion.^7,8^

SARS-CoV-2 belong to betacoronaviridae family, which includes mouse hepatitis coronavirus (MHV). The S1-NTD structure, described in the mouse hepatitis coronavirus (MHV), demonstrates a similar structural fold as the human galactose binding lectins (galectins).^9^ Galectins are carbohydrate-binding proteins, involved in many physiological functions, such as inflammation, immune responses, cell migration, autophagy and signalling.^10^ They are also involved immunogenicity, i.e. cell recognition, between human cells and infective pathogens; viruses, bacteria, and parasites.^11^

Prolectin – M is an orally administered polysaccharide. Polysaccharides competitively bind to the N-terminal tail of human galectin-3 through a proline isomerization.^12^ Galetin-3 (Gal-3) is 1 among the 15 galectins described in humans and also a ubiquitous human galectin expressed in various disease pathogenesis pathways.^13,14^ The objective of this trial is to demonstrate the feasibility of a large randomised controlled clinical trial, and a possible mechanism of action for galectin antagonists as treatment in COVID-19.

Viral culture studies of SARS-CoV-2^15^ indicate, that date of onset of symptoms and cycle threshold levels relate to infectivity; a cycle threshold of <25 is considered to be infectious.^16 17^ A cycle threshold (Ct) value is the number of replication cycles required for a signal of reverse transcription polymerase chain reaction (rRT-PCR) product, to cross a determined threshold.

Most rRT-PCR approved kits, report Ct values for the RNA dependent RNA polymerase (Rd/Rp), envelope (E) protein and nucleocapsid genes. All three genes are reported together for higher sensitivity. The droplet digital PCR measures expression of only nucleocapsid genes (N1&N2) as an absolute number in copies/uL.^18^ We hypothesized that blocking S1 NTD using a galectin antagonist can affect viral replication in nasopharyngeal, oropharyngeal, gastrointestinal tract, and potentially clear the virus from the person to make them non-infectious in a shorter time.^19^

For this pilot study we considered a rising number in the Ct values, along with a reduction in the number of copies/µL of the nucleocapsid gene as a measure of our endpoint. However, in future clinical trials Prolectin-M should be tested using an endpoint that measures the effect on time to recovery in days as well as the reduction of infectivity.^20,21^ A galectin antagonist could treat COVID-19 patients early in the disease pathogenesis and prevent spread of SARS-CoV-2 within the community.^22^

## Methods

### Trial Design and oversight

This investigation is a pilot and feasibility open-label randomized controlled trial. The trial protocol appears in supplement 1. A total of 10 subjects were identified between September 15^th^ and 19^th^, 2020. Study data was collected and managed using REDCap electronic data capture tools hosted at [Narayana Hrudayalaya Limited Bangalore].^23,24^

REDCap (Research Electronic Data Capture) is a secure, web-based software platform designed to support data capture for research studies, providing 1) an intuitive interface for validated data capture; 2) audit trails for tracking data manipulation and export procedures; 3) automated export procedures for seamless data downloads to common statistical packages; and 4) procedures for data integration and interoperability with external sources.

This study followed the Consolidated Standards of Reporting Trials (CONSORT) 2010 extension to randomized pilot and feasibility trials.^25^ The study was registered at clinicaltrials.gov (NCT04512027) and the Clinical Trials Registry of India (CTRI/2020/09/027833). Study protocol and all documents were reviewed by Institutional Ethics Committee – Narayana Health and approved (NHMEC Ref. No. S44/ 2020), on 31^st^ of August 2020. The study was conducted in compliance with the Declaration of Helsinki, the Good Clinical Practice guidelines, and local regulatory requirements.

Before enrolment and allocation to the study arms, informed consent for participation in the study was obtained in writing from all participants. The participant also signed a form, declaring that they received the correct information and gave informed consent for voluntary participation in the trial, before being randomized.

### Participants

This single center trial enrolled hospitalized, eligible, and consenting participants between the ages of 18 and 45. All patients had to have an instrumental diagnosis for COVID-19; a positive rRT-PCR for SARS-CoV-2 obtained from an outpatient collection of nasopharyngeal swabs. Other inclusion criteria were the presence of symptoms that were not older than 72 hours and an ability to provide consent to undergo repeated collection of throat and nasal swabs over the 7-day period. Samples were collected pre randomization, during drug administration on days 3 and 5 and 7.

Exclusion criteria included oxygen saturation at admission ≤96%, high temperature ≥100° F (≥37.5°C) not controlled on oral doses of acetaminophen, known history of diabetes on oral medications or insulin therapy or interleukin-6 levels ≥ 3times of laboratory reference range and / or significantly elevated levels of CRP, serum ferritin or d-dimer or a Lymphocyte / monocyte ratio ≤3 or neutrophil / lymphocyte ratio ≥5 or platelet count ≤150,000 cells per microliter. Previously tested positive and recovered for SARS-CoV-2. Also participants on any chronic medications for more than 4 weeks before randomization or active malignancy or having any co-morbidity that increases risk of rapid disease progression were excluded.

### Objectives

The primary aim was to evaluate the efficacy, safety and feasibility of administering Prolectin-M versus the SoC for 5 days. The primary endpoint was a change in absolute count of Nucleocapsid gene and a rising Ct value, estimated from serial samples of RNA extracted from a nasopharyngeal swab. The swab was collected in all participants on days 1 (day of randomisation), 3, 5, and 7. Clinical progression was estimated on a 7-point scale recorded on days 7, 21, and 28. A 2-point change was defined as clinical progression.

7-point severity score (ordinal scale):

1. Not hospitalized, no limitations on activities
2. Not hospitalized, limitation on activities.
3. Hospitalized, not requiring supplemental oxygen.
4. Hospitalized, requiring supplemental oxygen.
5. Hospitalized, on non-invasive ventilation or high flow oxygen devices.
6. Hospitalized, on invasive mechanical ventilation or extracorporeal membrane oxygenation (ECMO).
7. Death

### Randomization and Treatments

Participants were randomly assigned in a 1:1 ratio to receive a 4 gram tablet of Prolectin-M; a (1-6)-Alpha-D-Mannopyranose, and SoC (Treatment group) or SoC (Control group) via a web-based secure centralized system (REDCap). An independent statistician provided a computer-generated assignment randomization list and blocked with varying block sizes unknown to the investigators.

Prolectin-M was administered orally once every hour up to a maximum dose through the day of 40 grams or 10 tablets a day. The intention was to mimic the viral replication cycle of 8 – 10 hours^26^ and also to ensure that the participant is consuming the tablets during the day under supervision of a research nurse. Each subject was encouraged to keep the tablet in their mouth for 1-2 minutes before it dissolved and swallowed. During a mealtime, breakfast, lunch, tea, and dinner, the subject had to wait for 30 minutes after the last meal before taking the next tablet. This was to avoid any potential drop in blood glucose as the tablets could block absorption of carbohydrates consumed in the meal.

### Procedures

Patient follow up was maintained for 28 days from the day of randomisation. A sample was considered negative when no Ct value was determined, and no amplification curve was observed or if the Ct value was >29 for all three targets.

A nasopharyngeal / oropharyngeal swab was transported to the research laboratory for RNA extraction in a viral transport media (#MG20VTM-3, MagGenome). All RNA extractions were carried out using the QIAamp Viral RNA mini kit (# 52904, Qiagen) following the standard instructions as per the kit protocol.

For each blinded sample the Ct value for Rd/Rp and N, E gene, genes were reported.

#### rRT PCR

The extracted RNA was analysed on a rRT PCR platform: TRUPCR® SARS-CoV-2 RT qPCR KITV2 (#3B3043B BlackBio Biotech) SARS-CoV-2. To determine the efficiency of the PCR, Ct values obtained from a series of 5 template DNA dilutions of at least 3 different samples were graphed on the y-axis versus the log of the dilution on the x-axis.□The Ct values assumed by the following equation were employed to calculate the logarithm of the recombinant gene copy numbers from: Ct= slope × log (Gene Copy Number) +1 where I in the formula acts as the intercept of standard curve.^27^

#### Droplet Digital PCR (ddPCR)

The RNA was also analysed on the Droplet Digital PCR (BioRad, USA). This platform is FDA approved for emergency use authorization in COVID-19.^18^ The test is a partition-based endpoint single well RT-PCR test. The ddPCR was combined with rRT PCR because of its higher sensitivity and precision in low viral abundance samples and for the ability to provide absolute copy numbers and any resistance to inhibition often seen in rRT-PCR testing.

The analytic sensitivity was calculated as 0.260 cp/µl to 0.351 cp/µl (cp = copies) for genetic markers,□SARS-CoV-2□N1□and□N2□genes, and an internal control human RPP30 gene. The laboratory remained blind to treatment allocation throughout the analysis. On day 28, all participants were tested for serum antibodies against SARS-CoV-2 Immunoglobulin G (IgG) titres (values ≥1.00 as reactive) using VITROS® COVID-19 antibody tests.^28^

Adverse events were recorded from time of signature of informed consent and graded according to the Common Terminology Criteria for Adverse Events, version 5.0.^29^ Causality was assessed by the investigators for any serious adverse events.

### Statistical Analysis

Only 10 subjects were randomised and no formal sample estimation was done. Ct values and absolute copy numbers were compared using parametric, unpaired repeated t test with Welch’s correction or non-parametric Mann Whitney U test. A two-tailed, p<0.05, was considered statistically significant.

## Results

All 10 participants were included for final analysis. All participants consented and were randomised within 2 days of their symptom onset. All patients allocated to treatment arm received their medications. Demographic and baseline clinical characteristics are summarized in Table 1. 8/10 participants were female with a median (range) age of 27.5 (39, 25) years. In the control arm 3 patients did not have detectable viral RNA, when tested in the research lab. Table 2 shows changes that occurred over the follow up period. All treated participants showed rising Ct values from day 3. Difference in mean change in Ct values for the Rd/Rp & N genes, between treated and control group, was 16.41 (95% CI – 0.3527 to 32.48, p=0.047, Welch’s t test, two tailed). Difference in Ct values of E gene, 17.75 (95% CI;-0.1321 to 35.63, p = 0.05, unpaired t test with Welch’s correction, 2 tailed). Figures 2a & 2b show estimation plots for differences seen in Ct values. Figure 3 shows the change in copies/µL of the nucleocapsid (N1+N2) gene. A Mann Whitney test indicated that the drop in copy numbers among the treated participants was significant compared to the control group; U = 0.000, p<0.029. Individual patient wise change in absolute copy numbers is shown in supplementary figure panels.

**Table 1.** Baseline characteristics.

| Characteristic | Treated | Control |
| --- | --- | --- |
| Number | 5 | 5 |
| Age | 28.4 ± 3.195 | 29.1 ± 4.53 |
| Sex (female) | 4 | 4 |
| rRT PCR confirmed Sars-CoV-2 infection | 5 | 5 |
| Time from symptom onset to randomization (median) | 1.80 days | 2 days |
| WHO – CPS Score (0-10) = 4 | Score 2 (5) | Score 2 (5) |
| 7-point ordinal scale | 3 | 3 |
| Co – existing conditions | Nil | Nil |
| Laboratory Values |  |  |
| CRP, mg/L | 1.13 ± 0.80 | 4.93 ± 6.54 |
| D – Dimer, µg/L | 84.8 ± 78.25 | 95.60 ± 83.73 |
| Neutrophil Count | 63±15.24 | 53.82±9.19 |
| Lymphocyte count | 28.89±10.69 | 33.78±6.60 |
| Lymphocytes to neutrophil ratio | 0.691±0.557 | 0.617±0.17 |
| Lymphocyte to monocyte ratio | 0.41±0.91 | 0.38±0.23 |
| RDW | 14.58±1.90 | 16.69±2.89 |
| Hemoglobin | 13.24 ± 0.20 | 12.27±0.87 |
| Platelet count | 227±67.32 | 266.6±109.78 |
| ALT/ SGPT | 19 ± 8.98 | 16 ± 5.33 |
| AST / SGOT | 29 ± 8.04 | 25.6 ± 3.43 |
| Creatinine | 0.66 ± 0.11 | 0.85 ± 0.26 |
| Ferritin | 27.96 ± 36.69 | 13.30 ± 7.47 |
| LDH | 219 ± 39.96 | 197 ± 3 6.63 |
| Cycle thresholds | N = 5 | N = 2 |
| Rd/Rp + Nucleocapsid Gene | 21.306 | 19.549 |
| Envelope Gene | 23.126 | 22.0895 |

**Table 2:**
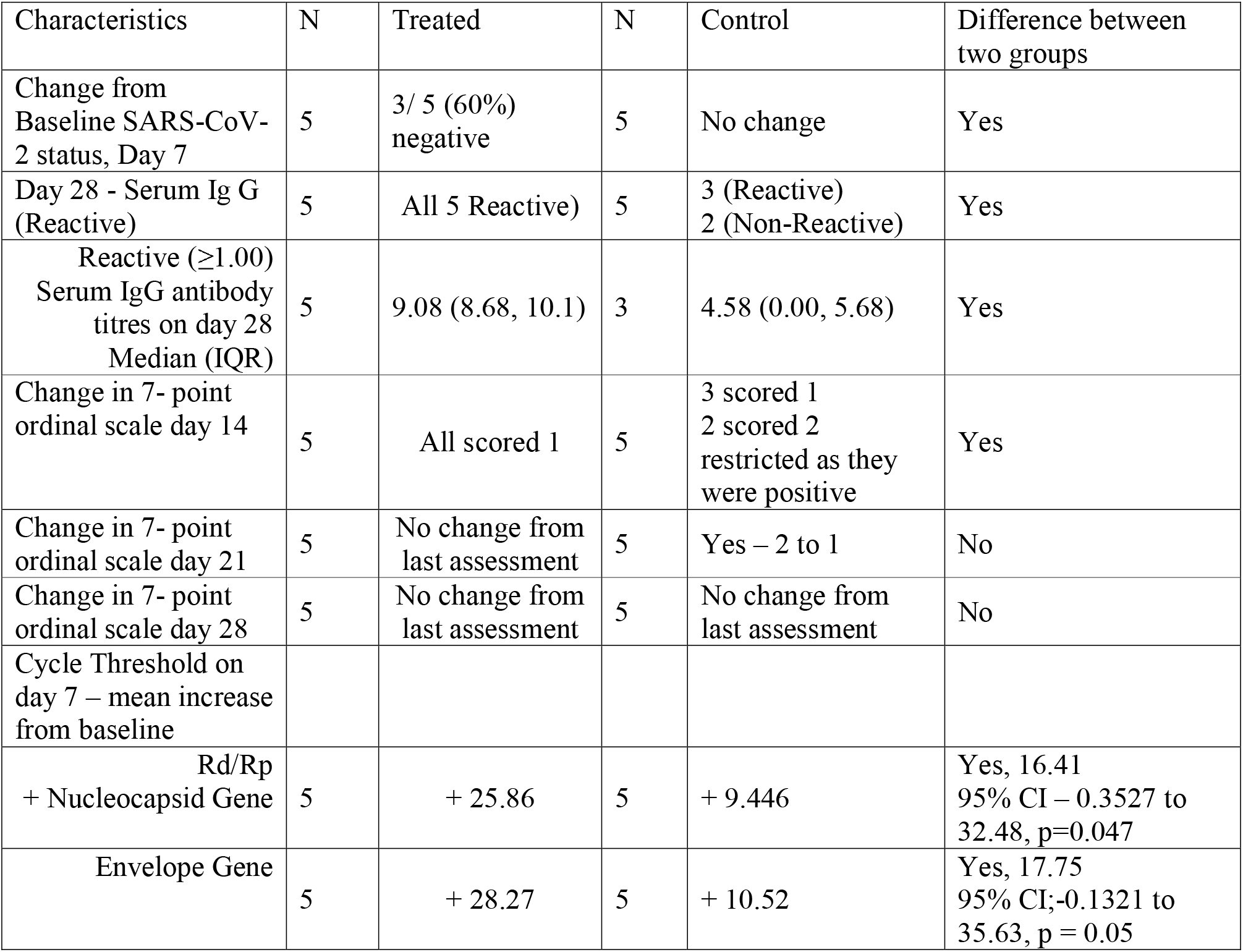
Follow up, changes seen after randomization.

| Characteristics | N | Treated | N | Control | Difference between two groups |
| --- | --- | --- | --- | --- | --- |
| Change from Baseline SARS-CoV-2 status, Day 7 | 5 | 3/ 5 (60%) negative | 5 | No change | Yes |
| Day 28 - Serum Ig G (Reactive) | 5 | All 5 Reactive) | 5 | 3 (Reactive)<br>2 (Non-Reactive) | Yes |
| Reactive ($\geq 1.00$)<br>Serum IgG antibody titres on day 28<br>Median (IQR) | 5 | 9.08 (8.68, 10.1) | 3 | 4.58 (0.00, 5.68) | Yes |
| Change in 7- point ordinal scale day 14 | 5 | All scored 1 | 5 | 3 scored 1<br>2 scored 2<br>restricted as they were positive | Yes |
| Change in 7- point ordinal scale day 21 | 5 | No change from last assessment | 5 | Yes – 2 to 1 | No |
| Change in 7- point ordinal scale day 28 | 5 | No change from last assessment | 5 | No change from last assessment | No |
| Cycle Threshold on day 7 – mean increase from baseline |  |  |  |  |  |
| Rd/Rp<br>+ Nucleocapsid Gene | 5 | + 25.86 | 5 | + 9.446 | Yes, 16.41<br>95% CI – 0.3527 to 32.48, p=0.047 |
| Envelope Gene | 5 | + 28.27 | 5 | + 10.52 | Yes, 17.75<br>95% CI;-0.1321 to 35.63, p = 0.05 |

**Figure 2a.**
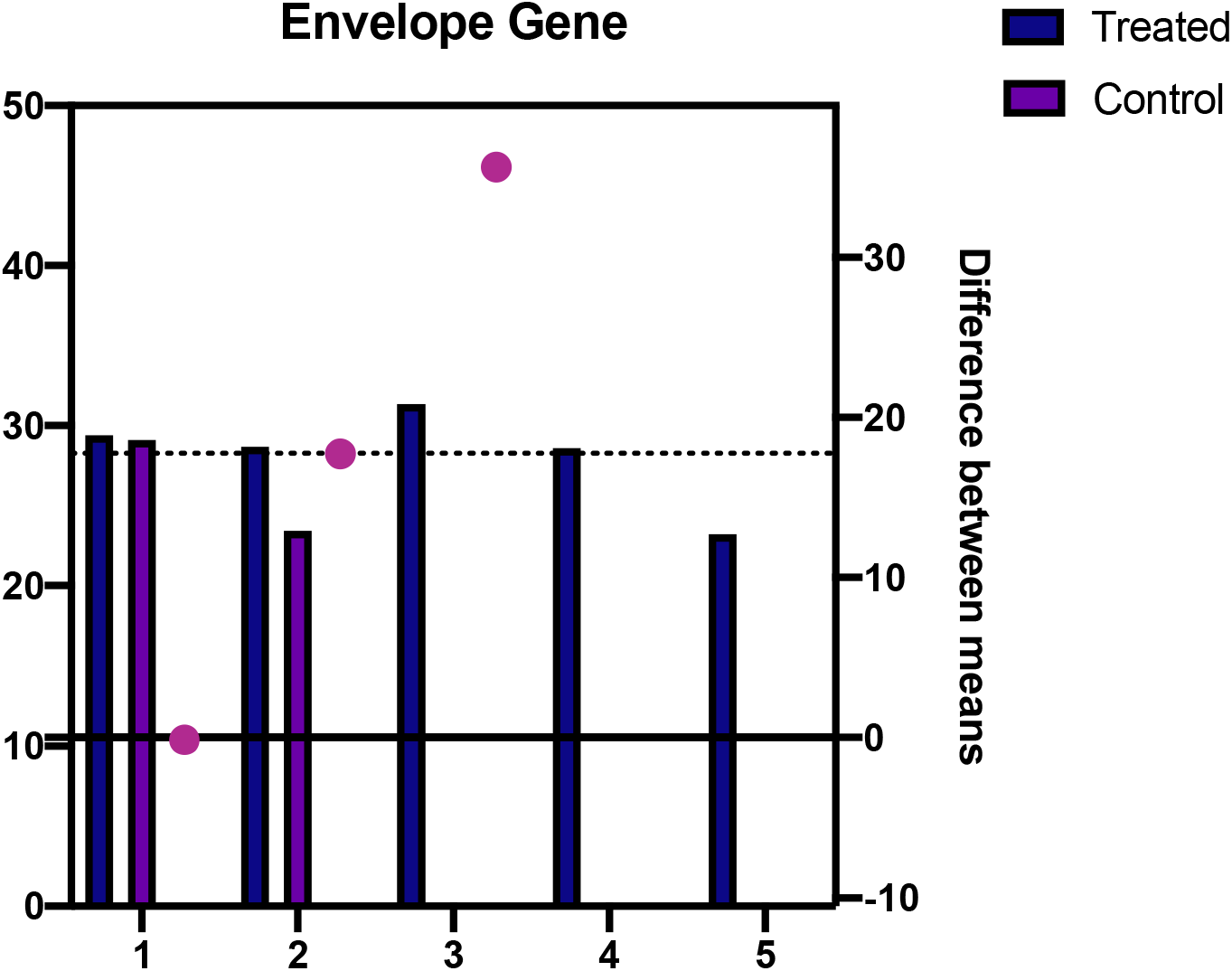
Change in Ct values over time – Rd/Rp+N gene

**Figure 2b.**
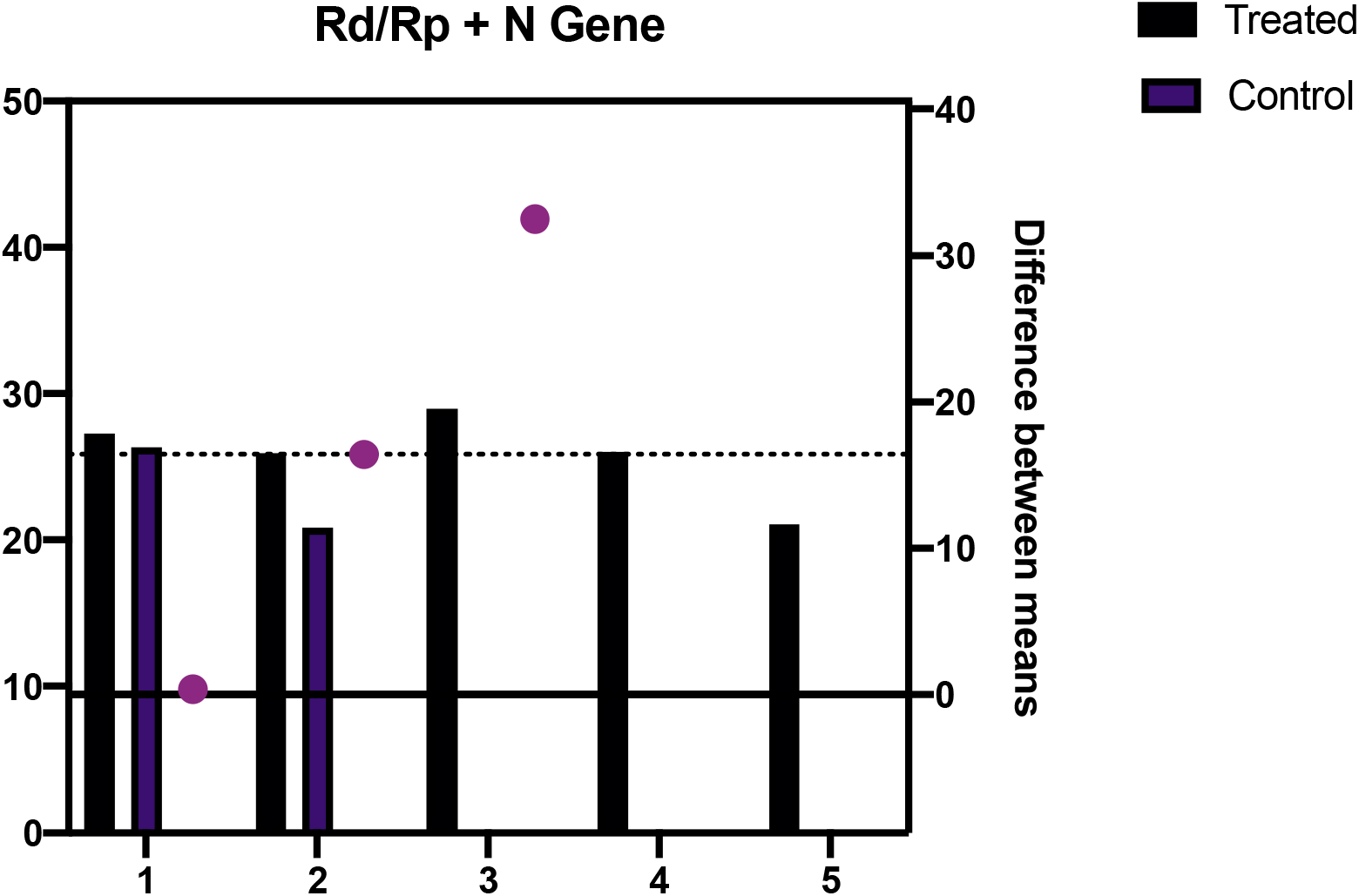
Change in Ct value over time – Envelope Gene

**Figure 3.**
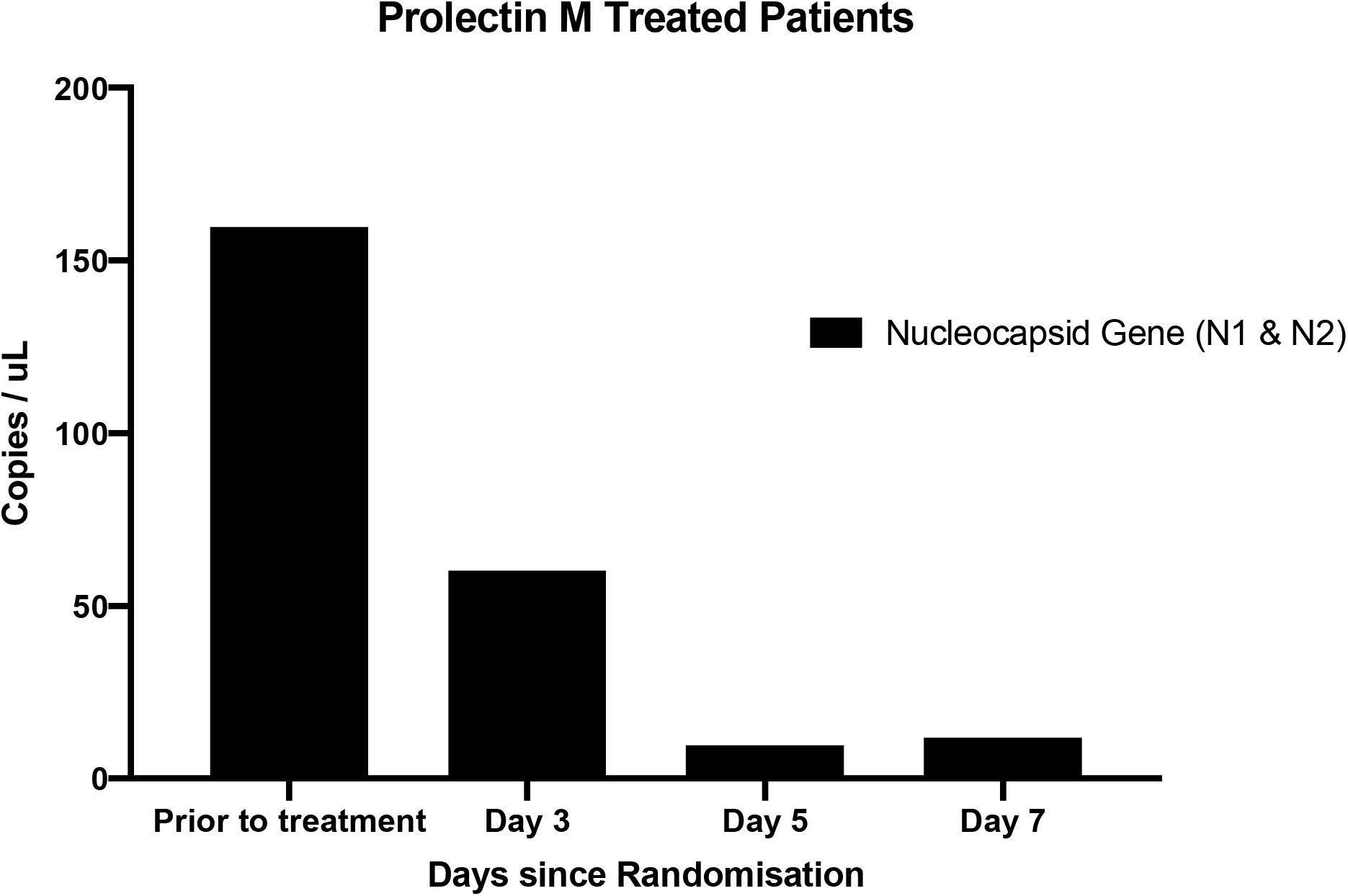
drop in absolute copy numbers of nucleocapsid gene over time – Treated group

**Figure 4.**
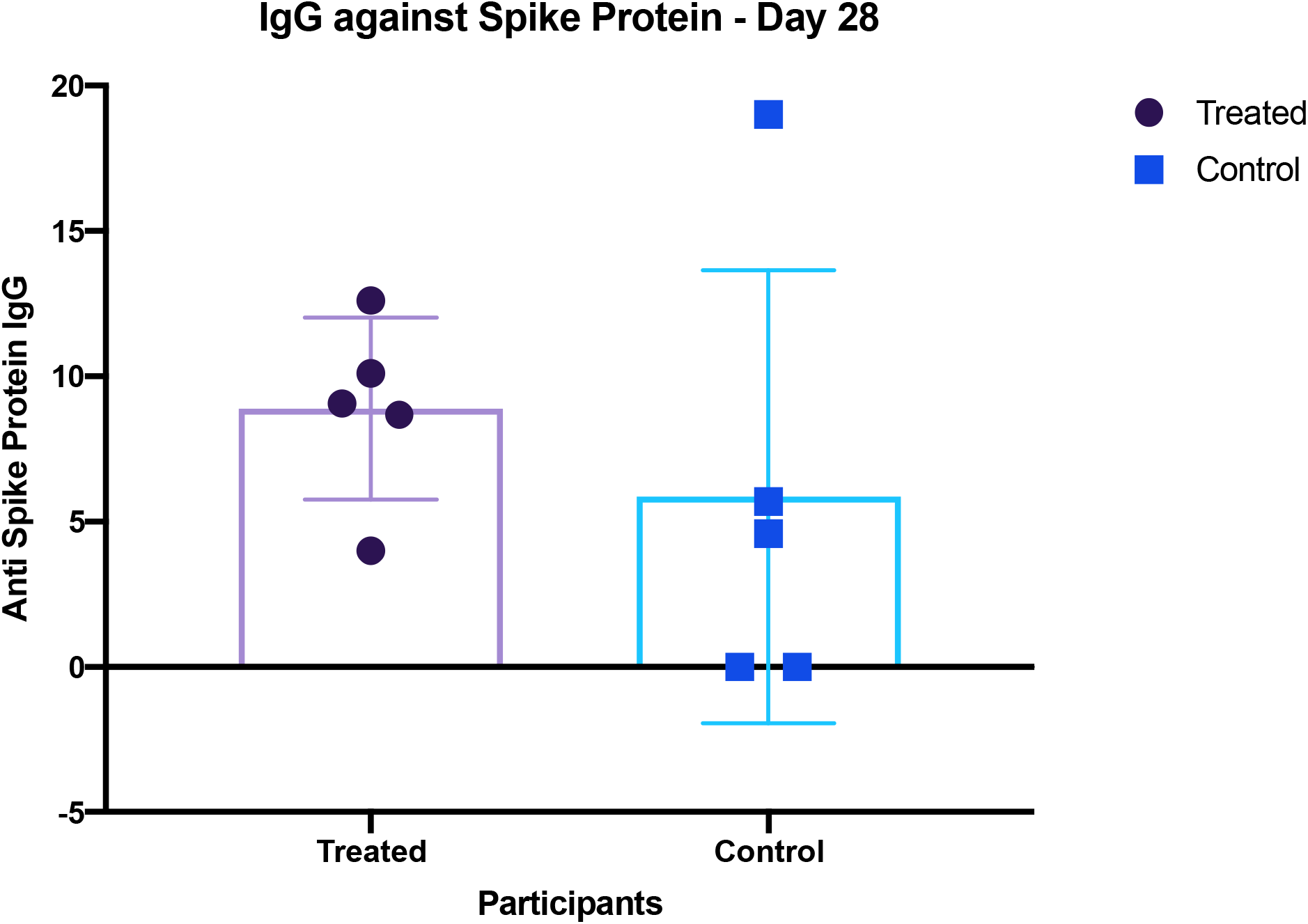
difference in IgG on day 28

rRT-PCR testing done in the clinic on day 1, 7, and 14 and had 3 participants (60%) turn negative by day 7 and all turned negative by day 14 and stayed negative until day 28. In the SoC group 2 participants had zero detectable viral loads at baseline, 2 participants tested negative on day 14, and the last participant tested remained positive on day 28.

None of the participants experienced any serious adverse event. One participant in the treatment arm had Grade 1 diarrheal episode (CTAE ver. 5.0). No intervention was need, but Prolectin-M was reduced to 5 tablets per day. All participants were fit for discharge on day 14 and with no restrictions except for 2 subjects in the control arm, who tested positive even on day 14. No one had re admission when followed up at day 21 and 28.

## Discussion

This is the first ever reported randomised controlled trial of a galectin antagonist against SARS-CoV-2. Blocking N terminal domain (NTD) of S1 subunit, present on spike protein, is seen to have significantly lowered viral gene expression. All five participants in treatment group demonstrated a rapid drop by day 3 in copies/µL of Nucleocapsid protein gene.

A higher Ct value correlates with lower risk of infectiousness, as reported from a cohort in England{ref}, There was rise in Ct >29, for genes, Rd/Rp, N and E genes in the treated group. However, participants in control group (001 & 007), remained potentially infective even on day 7 and day 14.

The detection of anti-spike protein IgG on day 28, is an expression of humoral response against the virus.^30^ The N protein is highly immunogenic compared to the S protein and in the absence of glycosylation sites on it results in N-specific neutralizing antibody production early in the stage of acute infection. All participants in the active arm of the trial had a reactive IgG. This trial achieved its primary objective of demonstrating the effect of Prolectin-M on lowering viral infectiousness. The method used to recruit and randomize patients, enabled safe administration of the oral drug which demonstrated an ability to block viral replication.

Strengths of this study are randomisation, concealed allocation, and one hundred percent compliance to treatment, and all procedures in the protocol. There was no incidence of any serious adverse events. A higher cut off value for either cycle threshold or copy numbers on ddPCR as an eligibility criteria could have demonstrated a more significant treatment effect. Hence future trials will need to use either a pre-determined lower (<16) baseline Ct value or a higher copy number for ddPCR.

Galectin antagonists can potentially prevent SARS-CoV-2 entry into human cells.^19^ A larger clinical trial will give us the evidence needed to understand its true clinical benefit. Due to the excellent safety profile of galectin antagonists, additional methods of delivery, such as an intravenous, or subcutaneous injection should be examined for a reduction in the number of systemically expressed copies of the virus. In addition, Prolectin-M could be very effective as a prophylactic to stop the community spread of the disease. The results from this pilot and feasibility trial sets the stage for a definitive large randomised controlled clinical trial using polysaccharides as galectin antagonists in order to study their potential role as a Post Infection Immunisation (PII) which could lower the basic reproduction number (Ro) of the disease by lowering the levels of contagiousness and transmissibility.

## Data Availability

This is a pilot and feasibility trial. The data is available in a secure database and is locked within a REDCap electronic database.

## Acknowledgements

There are no acknowledgements to be mentioned.

## Conflict of Interest

The corresponding author was the listed Principal Investigator for the trial at Narayana Hrudayalaya Limited and is the Group Head of Clinical Research. His department has received remuneration from the company – Pharmalectin Inc, towards conducting the clinical trials at the center.

David Platt is the CEO of Pharmalectin Inc. However he or his team did not have any access to the data, during the trial and after completion. They did not involve in the statistical analysis and only contributed as a co-author in this manuscript.

Other Co – authors have no specific conflict of interest to declare.

---

**Supplementary Figure 1 (a – e).**
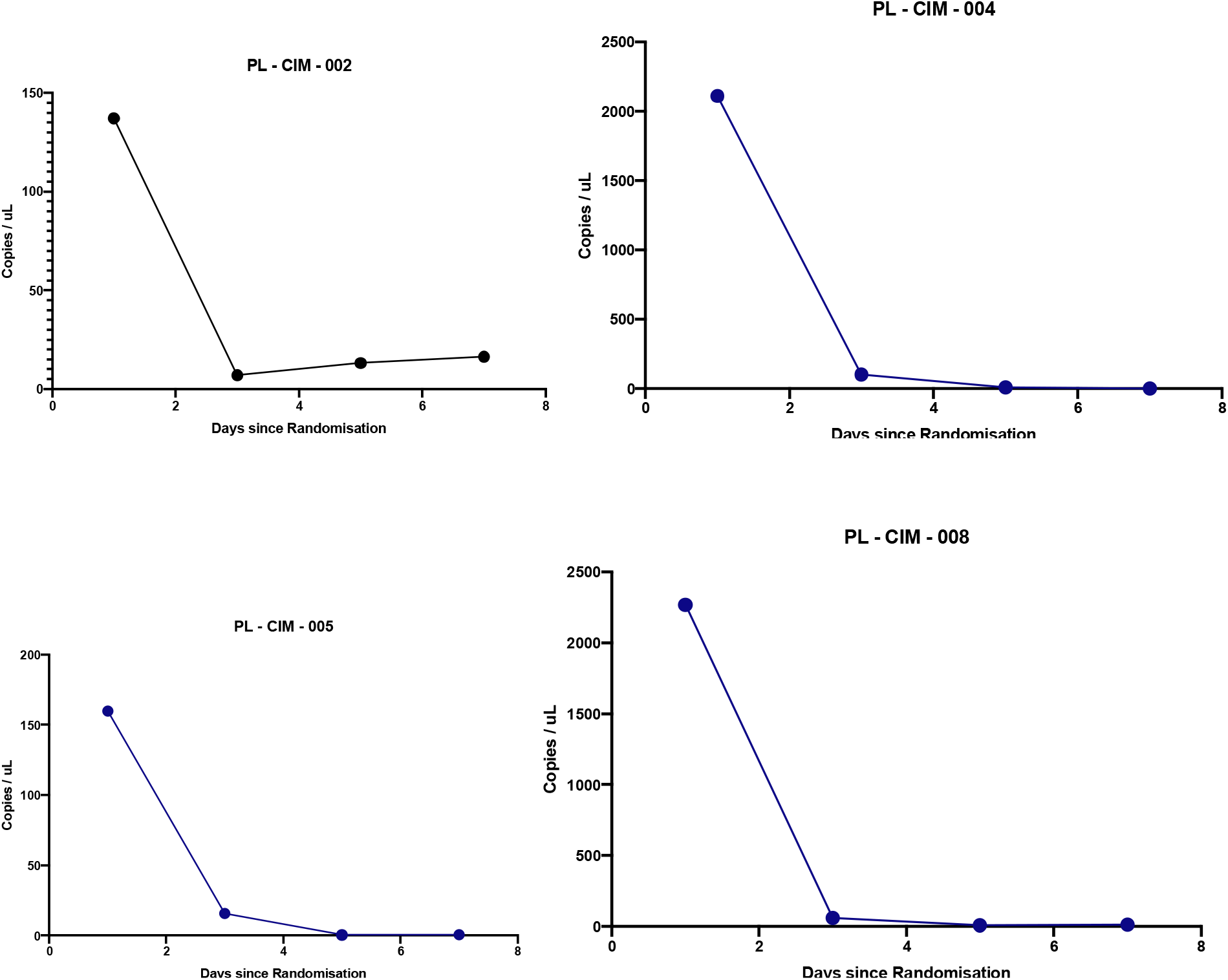
Nucleocapsid gene expression in Treated group participant

**Supplementary Figure 2.**
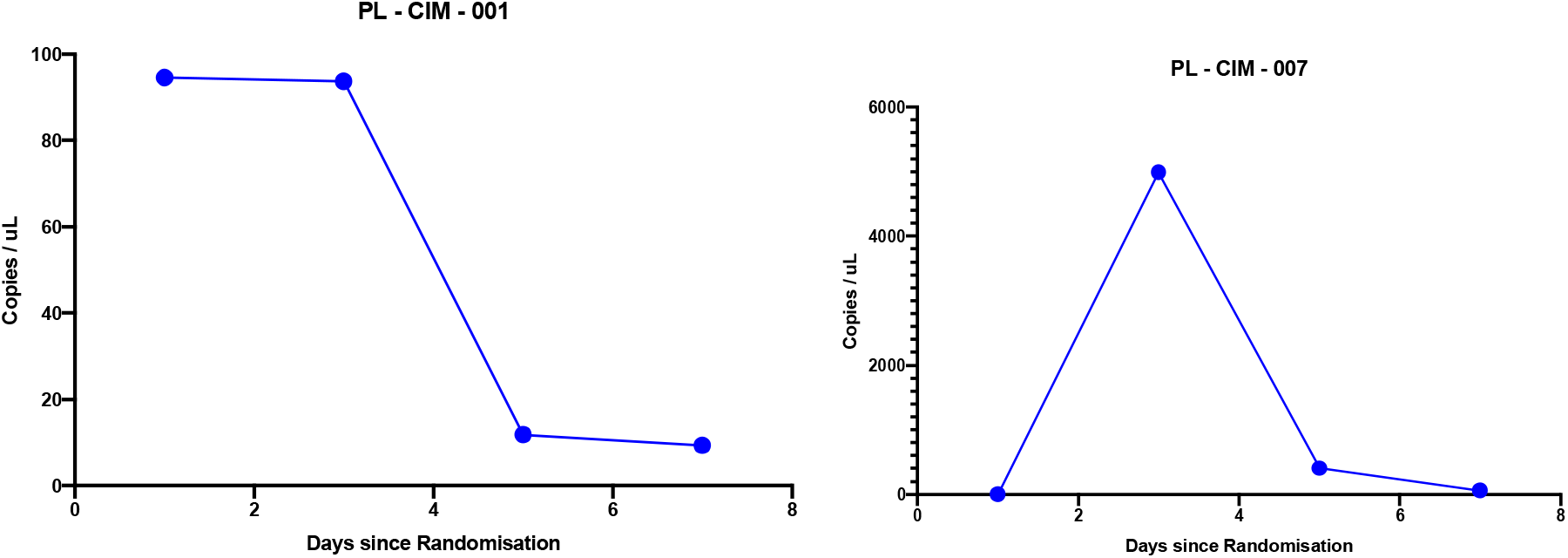
Nucleocapsid gene expression in Control group participants

